# Association of Sedentary Time with Hypertension and Mediating Effect of Uric Acid to High-Density Lipoprotein Ratio: an Analysis Based on NHANES 2017-2020 Data

**DOI:** 10.1101/2025.04.26.25326500

**Authors:** Guangling Li, Jing Tao, Jie Fu, Chao Wang

**Affiliations:** Department of Anesthesiology, Affiliated Hospital of Jiangnan University,No.1000,Hefeng Road, Wuxi, 214125, China; Department of Anesthesiology, the First Affiliated Hospital of Bengbu Medical University, Bengbu, China; Department of Anesthesiology,The Affiliated Stomatological Hospital of Xuzhou Medical University,Xuzhou, China; Department of Gastrointestinal Surgery, Affiliated Hospital of Jiangnan University,No.1000,Hefeng Road, Wuxi, 214125, China

**Author notes:** Correspondence: Chao Wang, 4.Department of Gastrointestinal Surgery, Affiliated Hospital of Jiangnan University,No.1000,Hefeng Road, Wuxi, 214125, China.

**Keywords:** Sedentary time, Hypertension, Uric acid to high-density lipoprotein ratio, Mediating effect, NHANES

## Abstract

**BACKGROUND:** Sedentary behavior has been recognized as an important public health problem, but the specific mechanisms linking it to hypertension have not been fully elucidated.

**OBJECTIVE:** To explore the association between sedentary time and hypertension and to assess the mediating role of uric acid to high-density lipoprotein ratio (UHR) in the U.S. adult population.

**METHODS:** Data from the 2017-2020 National Health and Nutrition Examination Survey (NHANES) were utilized. Weighted multiple logistic regression analysis was used to assess the association between sedentary time and hypertension, and the role of UHR was assessed by mediation effect analysis.

**RESULTS:** Sedentary time was found to be significantly and positively associated with hypertension. This association remained significant even after adjusting for sex, age, race, socioeconomic status, and other confounders.The UHR mediated the association between sedentary time and hypertension with a mediation ratio of 0.123 (95% CI: 0.04 to 0.43, p = 0.002), indicating that approximately 12.3% of the total effect was mediated by the UHR. All analyses were adjusted for confounding variables such as sex, age, race, and BMI. These results suggest a positive association between sedentary time and hypertension, partially mediated through UHR.

**CONCLUSION:** In US adults, increased sedentary time was found to be significantly associated with hypertension, and this association was partially mediated by UHR. Further studies are needed to validate this.

## 1. Introduction

In recent years, sedentary behavior has emerged as a significant public health concern globally, particularly due to its association with various chronic diseases. Among these, hypertension stands out as a prevalent condition that is closely linked to sedentary lifestyles. Research indicates that a sedentary lifestyle is a common risk factor for hypertension, with studies revealing that individuals who engage in prolonged sitting are at a higher risk of developing elevated blood pressure levels [1]; [2], [3]. For instance, a study highlighted that 36% of participants were categorized as sedentary, underscoring the widespread nature of this issue [1]. Furthermore, sedentary behavior has been shown to independently increase the risk of obesity, type 2 diabetes, and hypertension, suggesting that the relationship between sitting time and high blood pressure is multifaceted [4]; [5].

Despite the established correlation between sedentary behavior and hypertension, the precise mechanisms through which prolonged sitting influences blood pressure remain inadequately understood. One proposed mechanism involves the reduction of energy expenditure associated with sedentary activities, which can lead to metabolic dysregulation [6]. Specifically, prolonged sitting may result in decreased lipoprotein lipase activity, an enzyme critical for lipid metabolism, thus contributing to elevated glucose and lipid levels in the bloodstream, which are risk factors for cardiovascular diseases [4]. Additionally, sedentary behavior may displace opportunities for engaging in light-intensity physical activities, further exacerbating metabolic syndrome and hypertension [7].

Moreover, the impact of sedentary behavior on cardiovascular health is compounded by its association with obesity. Studies have shown that obesity can mediate the relationship between sitting time and hypertension, indicating that excess body weight may amplify the adverse effects of prolonged sitting ; [2]. Assuch, interventions aimed at reducing sedentary time, such as promoting regular physical activity or implementing sit-stand workstations, have been suggested as potential strategies to mitigate hypertension risk [8]; [9]. However, further research is necessary to elucidate the underlying biological mechanisms and to develop effective public health strategies that address this growing concern.

In conclusion, while the link between sedentary behavior and hypertension is well-documented, the exact pathways through which sitting time affects blood pressure are still being explored. Understanding these mechanisms is crucial for developing targeted interventions to combat the rising prevalence of hypertension associated with sedentary lifestyles.

## 2. Methods

### 2.1 Study population

We analyzed records from the National Health and Nutrition Examination Survey (NHANES) database covering the period from 2017 to 2020. This survey assesses nutrition and health, and its protocol was approved by the NCHS Ethics Review Board. All participants provided written informed consent. Since the data were obtained from the official NHANES website (https://www.cdc.gov/nchs/nhanes/index.htm),no further ethical approval was necessary.Out of the initial 49,693 participants, exclusions were as follows: (1)individuals younger than 20 years old; (2) those without MHR data or identified as outliers; (3) those without ST data or recording less than 30 minutes; (4) those missing physician’s documented hypertension diagnosis or blood pressure readings. Consequently, 24,614 participants qualified for the study. The recruitment process is illustrated in Figure 1.

**Fig 1.**
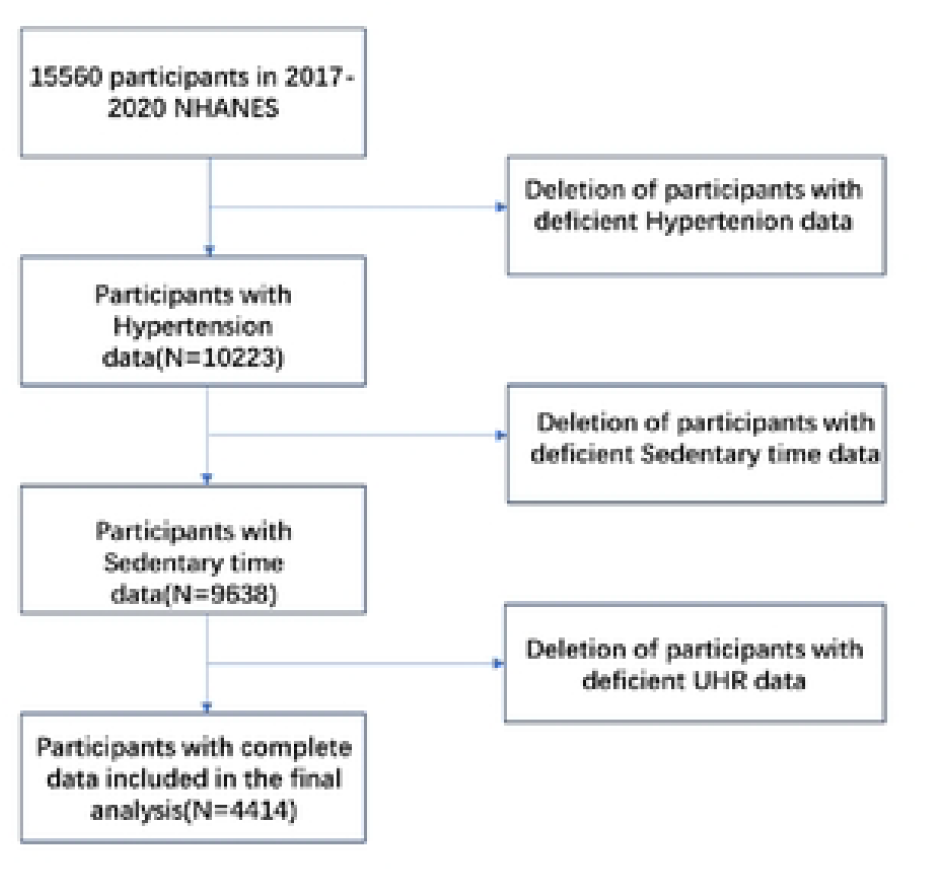
Flowchart showing the participants enrollment procedure

**Figure 2.**
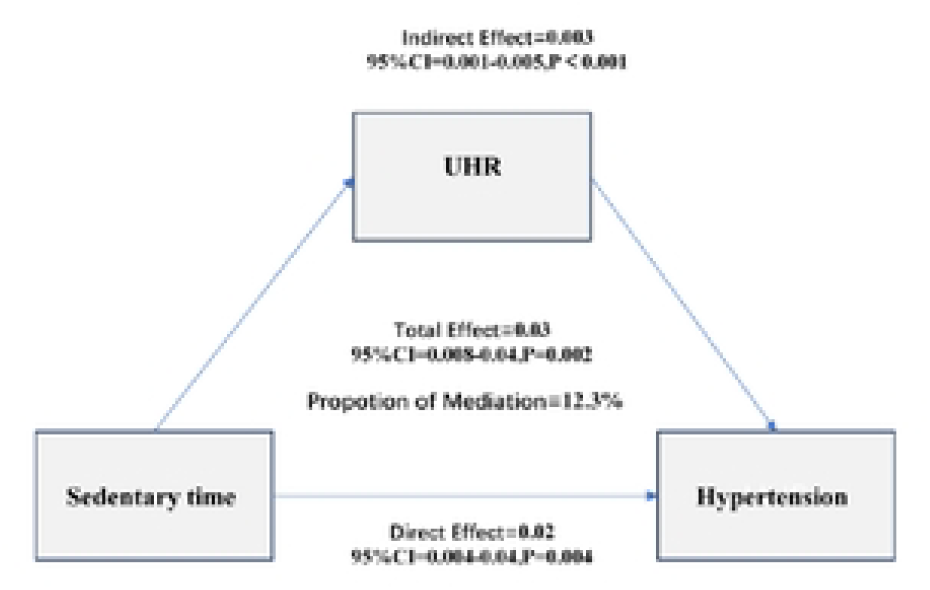
Mediation effect of UHR on the association between sedentary time and hypertension.

### 2.2. Study variables

Hypertension diagnosis was confirmed if any of the following criteria were met: average systolic blood pressure ≥140 mmHg, average diastolic blood pressure ≥90 mmHg, a physician’s diagnosis, or the use of antihypertensive medication.

HDL and uric acid were measured from blood samples drawn from participants in a fasting state in the morning. The steps for HDL measurement were as follows: A magnesium sulfate/dextran solution was added to the sample, forming water-soluble complexes with non-HDL cholesterol, which did not react with the measurement reagents in subsequent steps. Then, by adding polyethylene glycol esterase, HDL cholesterol esters were converted to HDL cholesterol. The hydrogen peroxide generated in this reaction reacted with 4-aminoantipyrine and HSDA to form a purple or blue dye. Finally, laboratory researchers determined HDL levels by photometric measurement at 600nm. The steps for UA measurement were as follows: Serum uric acid concentration was measured using the timed endpoint method with a DxC800 automated chemical analyzer. Uric acid was oxidized by uricase to produce allantoin and hydrogen peroxide. Hydrogen peroxide reacted with 4-aminoantipyrine (4-AAP) and 3,5-dichloro-2-hydroxybenzenesulfonate (DCHBS) in a peroxidase-catalyzed reaction to produce a colored product, which was then measured photometrically at 520 nm to determine uric acid levels. Then, UHR (%) was then calculated by dividing UA (mg/dL) by HDL (mg/dL) and multiplying by 100[10].

#### Sedentary time

Participant responses to the Global Physical Activity Questionnaire (GPAQ) were used to measure total sitting time each day, which has previously been validated to measure daily physical activity and sedentary behavior [11]. An in-person interview consisted of the following questions: “On a typical day, how much time do you usually spend sitting at school, at home, getting to and from places, or with friends, including time spent sitting at a desk, traveling in a car or bus, reading, playing cards, watching television, or using a computer?”

### 2.3 Covariate assessment

To accurately identify the factors influencing our study outcomes, weconsidered a range of covariates from the NHANES website (www.cdc.gov/nchs/nhanes/), which provides a detailed description of the datacollection process. These covariates include: (1) demographics data:age, gender, race, education level, and family income to poverty rate(PIR). (2) examination, laboratory, dietary and questionnaire data: body mass index (BMI), fasting blood glucose (FBG), energy intake, drinking,smoking, disease status, and the use of anti-hypertensive drugs.

### 2.4. Statistical analysis

Continuous variables were expressed as means with standard errors (SE), and categorical variables were presented as frequencies and percentages. Differences between groups (categorized by sedentary behavior) were assessed using independent t-tests for continuous variables and chi-square tests for categorical variables. Statistical significance was determined at a two-sided P-value < 0.05.To evaluate the association between sedentary time and hypertension, logistic regression models were employed. Three models were constructed: a non-adjusted model, an adjusted model (Adjust I), and a fully adjusted model (Adjust II). Adjust I included adjustments for gender, age, race, education, marital status, and poverty ratio (PIR), while Adjust II further adjusted for BMI, alcohol consumption, diabetes, coronary heart disease (CHD), and smoking status. Odds ratios (OR) and 95% confidence intervals (CI) were calculated for sedentary quartiles, with the lowest quartile serving as the reference group. A trend test was performed to assess the dose-response relationship between sedentary behavior and hypertension.Causal mediation analysis was conducted to explore the mediating role of UHR (uric acid to HDL ratio) in the relationship between sedentary behavior and hypertension. A generalized linear model with a probit link function was used, adjusting for gender, age, race, education, marital status, PIR, BMI, alcohol consumption, diabetes, CHD, and smoking status.

Nonparametric bootstrap resampling (1,000 iterations) was applied to estimate confidence intervals for the mediation effect. The total effect, direct effect, and mediation effect were reported, along with the proportion of the total effect mediated by UHR. Statistical significance was set at P < 0.05 for all analyses.

All statistical analyses were conducted using EmpowerStats (www.empowerstats.com) and R software.

## 3. Results

## 3.1. Characteristics of study population

Baseline characteristics of the study population (n=4411) are summarized in Table 1. Hypertensive patients (n=1513) had significantly higher sedentary time (6.39 ± 0.10 hours versus 5.89 ± 0.13 hours, P<0.001), UHR (12.18 ± 0.14 versus 10.28 ± 0.12, P<0.001), and uric acid (5.75 ± 0.05 versus 5.16 ± 0.04, P<0.001), while the hypertensive group had significantly lower HDL levels (52.52 ± 0.79 versus 55.27 ± 0.47, P<0.001). The hypertensive group was also older (55.98 ± 0.89 versus 41.99 ± 0.58 years, P<0.001) and had a higher prevalence of diabetes (22.75% versus 4.31%, P<0.001) and coronary heart disease (9.46% versus 1.08%, P<0.001). In addition, the hypertension group had a higher proportion of participants with a BMI ≥30 kg/m^2^ (57.49% versus 37.49%, P<0.001) and a lower proportion of individuals who had never smoked (17.57% versus 23.49%, P=0.003). There was no significant difference in poverty rates between the two groups (P=0.805).

**TABLE 1.**
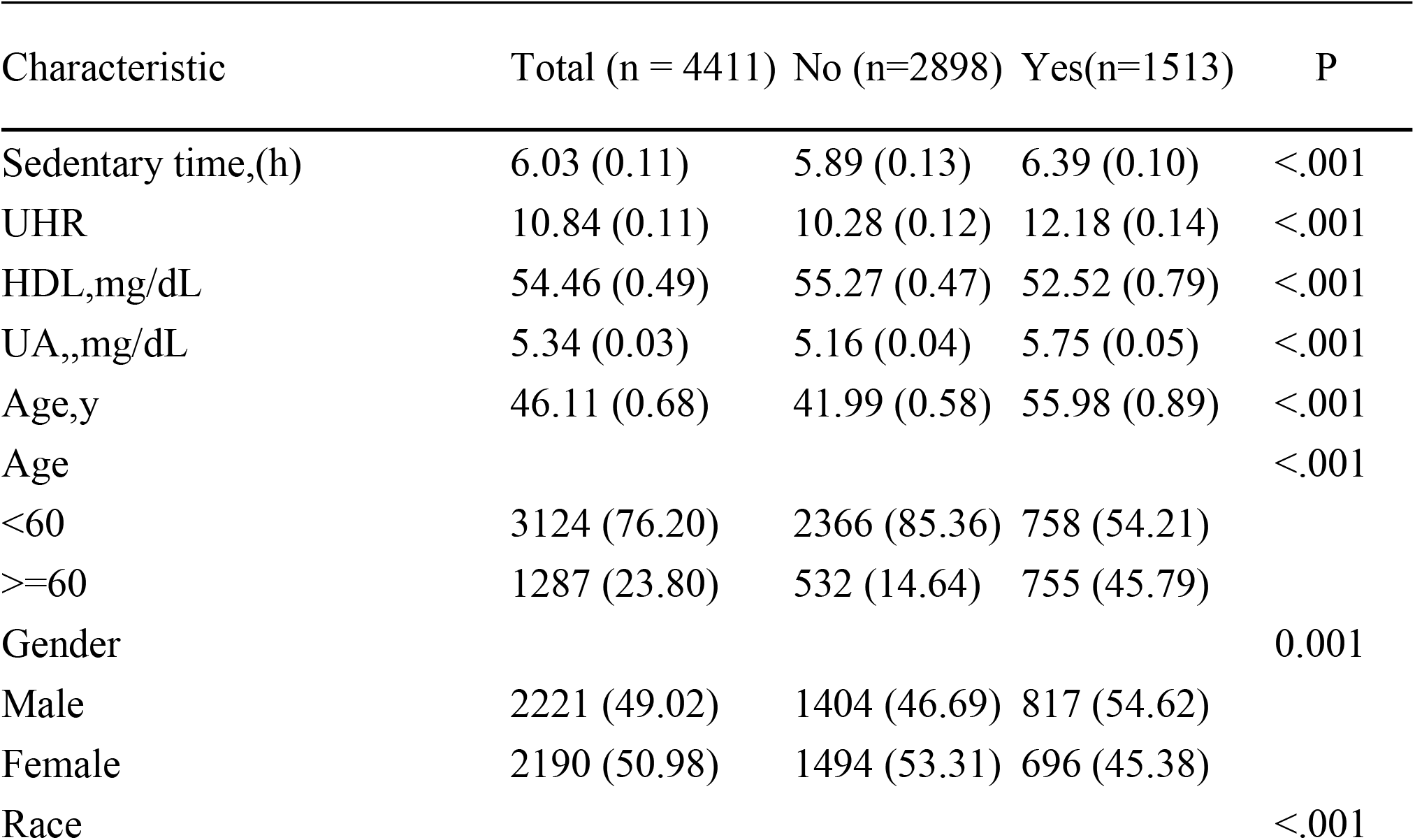
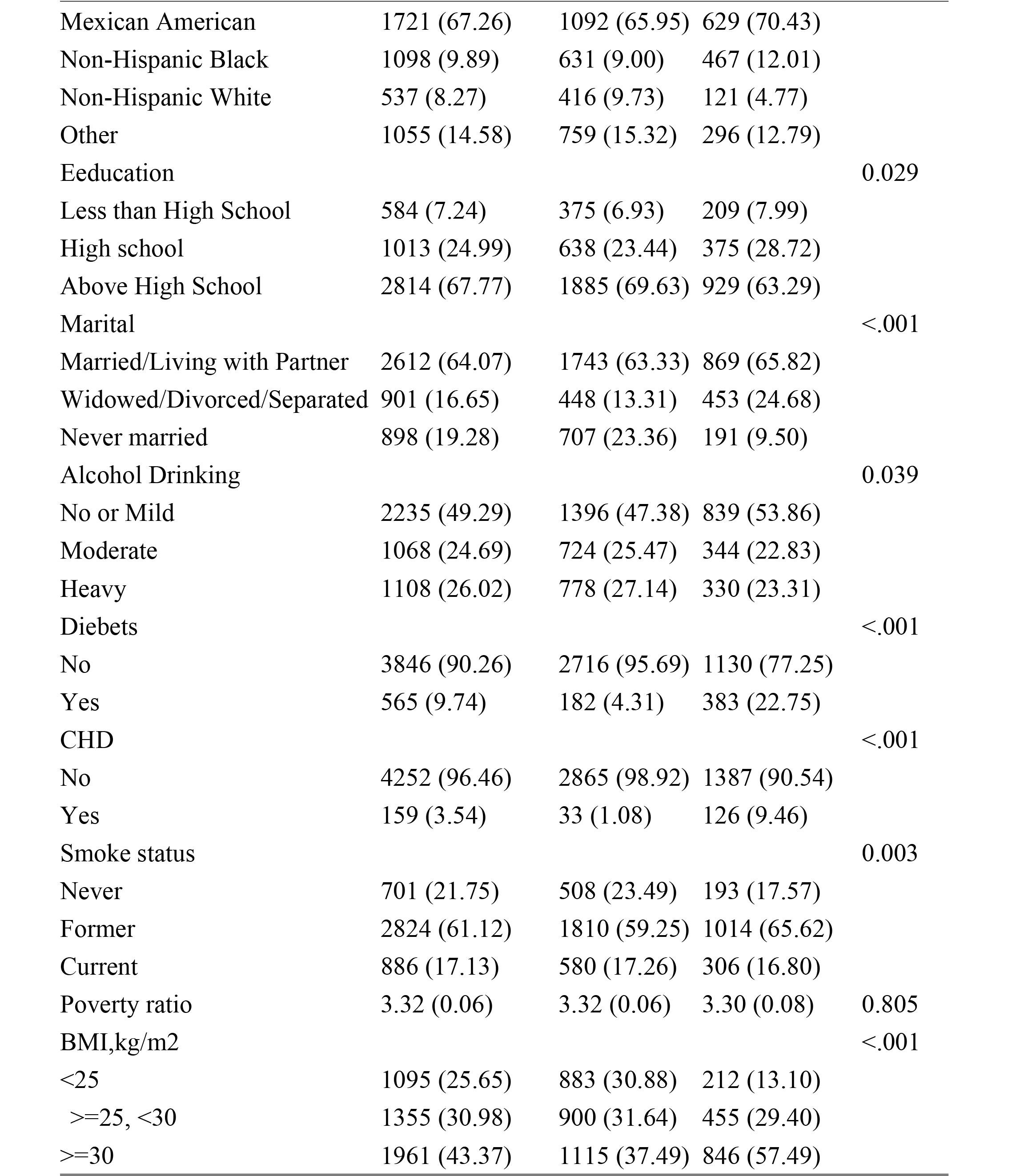
Baseline characteristics of participants in NHANES 2017−2020. Baseline characteristics of Participants. All proportions, means, and SEs are weighted estimates of the US population characteristics, taking into account the complex sampling design of the National Health and Nutrition Examination Survey. UA,Uric acid, HDL,High density lipoprotein,UHR,Uric acid to high-density lipoprotein ratio,CHD,Coronary heart disease;BMI, body mass index (calculated as weight in kilograms divided by height in meters squared).

### 3.2 Association between sedentary time and hypertension

In all models, sedentary time was positively associated with hypertension. In the unadjusted model, the odds ratio (OR) for hypertension was 1.04 (95% CI: 1.03-1.06, p<0.0001) for each hour increase in sedentary time. After adjusting for demographic and socioeconomic factors (model I), the OR increased to 1.05 (95% CI: 1.03-1.08, P<0.0001). In the fully adjusted model (Model II), which accounted for additional confounders such as BMI, smoking, alcohol consumption, diabetes, and coronary heart disease, the association remained significant with an OR of 1.03 (95% CI: 1.01-1.05, P=0.0125). In the sedentary quartile, the highest quartile (Q4) showed a significantly increased risk of hypertension (OR=1.36, 95% CI: 1.08-1.73, P=0.0101) compared to the lowest quartile (Q1).

**Table 2.** Association Between Sedentary time and Hypertension in Different Models.

| Exposure | Crude | Model I | Model II |
| --- | --- | --- | --- |
| Sedentary time continuous | 1.04 (1.03, 1.06) <0.0001 | 1.05 (1.03, 1.08) <0.0001 | 1.03 (1.01, 1.05) 0.0125 |
| Sedentary time quartile |  |  |  |
| Q1 | 1.0 | 1.0 | 1.0 |
| Q2 | 1.29 (1.05, 1.57) 0.0148 | 1.17 (0.93, 1.46) 0.1727 | 1.09 (0.86, 1.37) 0.4880 |
| Q3 | 1.56 (1.27, 1.90) <0.0001 | 1.39 (1.10, 1.74) 0.0048 | 1.26 (0.99, 1.59) 0.0584 |
| Q4 | 1.62 (1.33, 1.97) <0.0001 | 1.70 (1.36, 2.13) <0.0001 | 1.36 (1.08, 1.73) 0.0101 |
| P for trend | <.001 | <.001 | <.001 |

Crude model adjust for: None
Model I model adjust for: Gender; Age; Race; Education; Marital; PIR
Model II model adjust for: Gender; Age; Race; Education; Marital; PIR ;BMI;Alcohol Drinking;Smoke status;CHD

### 3.3 Subgroup analyses

Subgroup analyses showed that the association between sedentary time and hypertension was consistent across most demographic and clinical subgroups, with no significant interactions between sex (P=0.394), race (P=0.705), marital status (P=0.702), or diabetes status (P=0.957). However, smoking status (P=0.034) showed a significant interaction, with the strongest association for current smokers (OR=1.14, 95% CI: 1.08-1.21, P <0.001). In addition, participants with a BMI between 25-30 kg/m^2^ had a slightly stronger association with hypertension (OR=1.05, 95% CI: 1.01-1.10, P=0.029), compared with participants with a BMI <25 kg/m^2^ or ≥30 kg/m^2^. Table 3. Associations between sedentary time and hypertension stratified by selected demographic and clinical factors, NHANES, 2017–2020

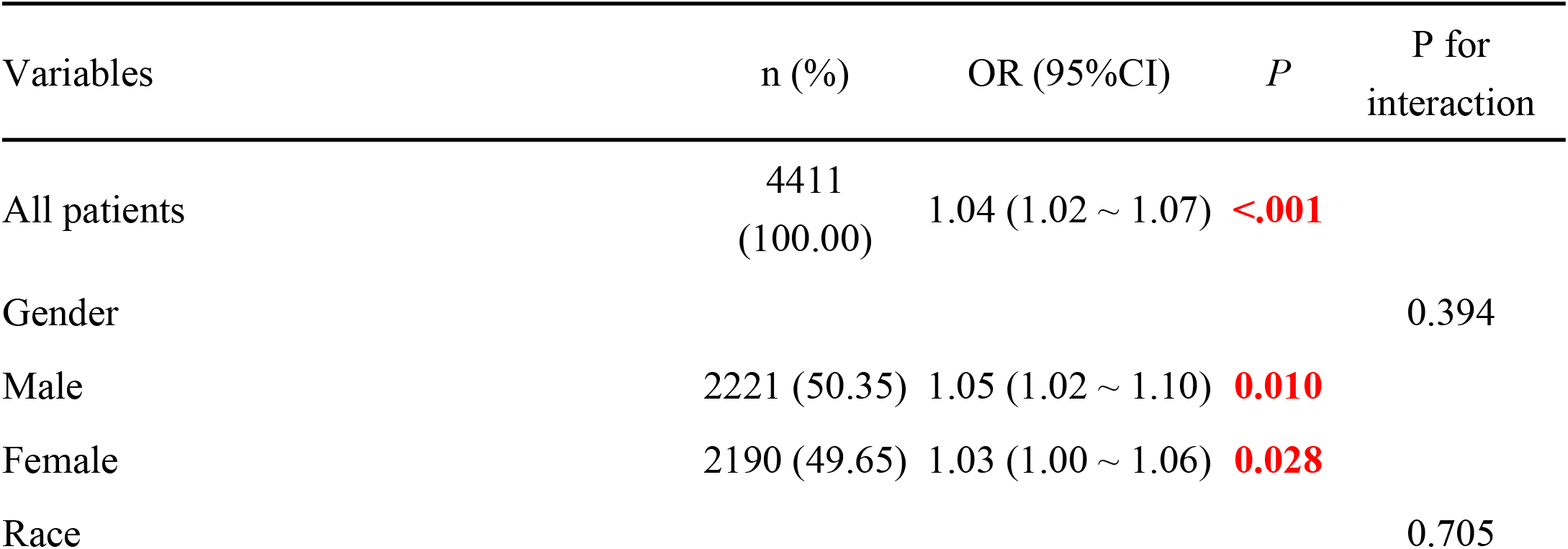
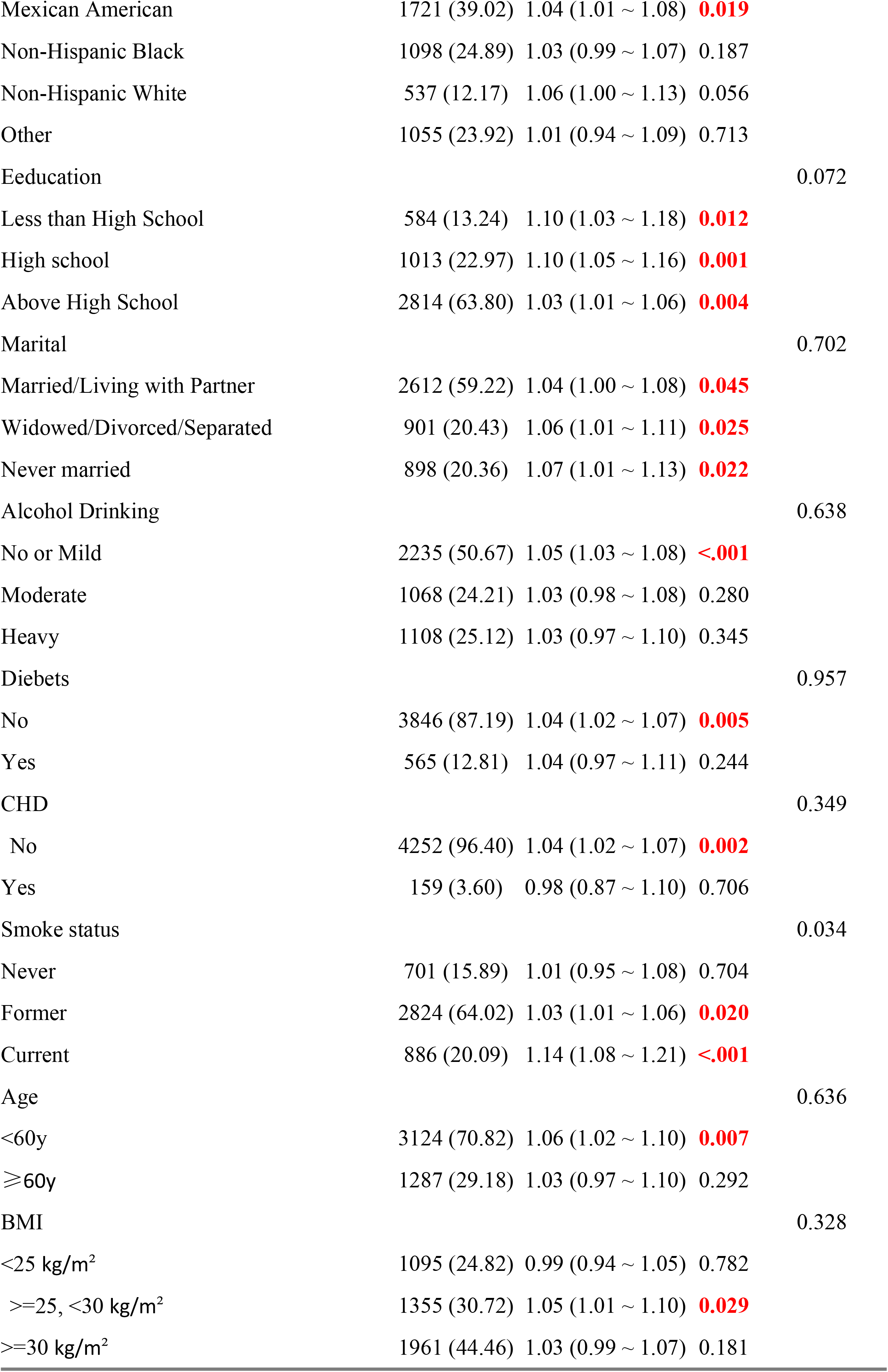
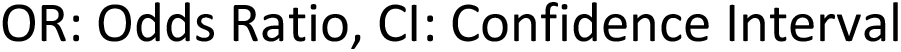

### 3.4 Association between sedentary time and UHR

Sedentary time was also positively associated with UHR in all models. In the unadjusted model, UHR increased significantly for each hour increase in sedentary time (β=0.1, 95% CI: 0.1-0.2, p <0.001). After adjusting for demographic and socioeconomic factors (Model I: β=0.2, 95% CI: 0.1-0.2, P <0.001) and further adjusting for lifestyle and clinical variables (Model II: β=0.1, 95% CI: 0.0-0.1, P <0.001), this association remained robust. In the sedentary quartile, the highest quartile (Q4) showed the greatest increase in UHR (β=0.73, 95% CI: 0.34-1.11, P=0.0002) compared to the lowest quartile (Q1).

**Table 4.** Association Between Sedentary Time and UHR in Different Models, NHANES, 2017– 2020.

| Exposure | Non-adjusted | Model I | Model II |
| --- | --- | --- | --- |
| SEDENTARY | 0.1 (0.1, 0.2) <0.001 | 0.2 (0.1, 0.2) <0.001 | 0.1 (0.0, 0.1) <0.001 |
| SEDENTARY quartile |  |  |  |
| Q1 | 0 | 0 | 0 |
| Q2 | 0.49 (0.03, 0.95) 0.0370 | 0.55 (0.13, 0.96) 0.0099 | 0.21 (-0.17, 0.59) 0.2810 |
| Q3 | 0.41 (-0.05, 0.87) 0.0783 | 0.75 (0.33, 1.17) 0.0005 | 0.26 (-0.12, 0.65) 0.1771 |
| Q4 | 1.03 (0.58, 1.48) <0.0001 | 1.48 (1.06, 1.90) <0.0001 | 0.73 (0.34, 1.11) 0.0002 |
| P for trend | <.001 | <.001 | <.001 |

Non-adjusted model adjust for: None
Model I model adjust for: Gender; Age; Race; Education; Marital; PIR
Model II model adjust for: Gender; Age; Race; Education; Marital; PIR ;BMI;Alcohol Drinking;Smoke status;CHD

### 3.5 Causal mediation analysis

Causal mediation analysis showed that UHR partially mediated the relationship between sedentary time and hypertension. The total effect of sedentary time on hypertension was significant (β=0.026, 95% CI: 0.008-0.045, P=0.0020), with a mean mediating effect of 0.003 (95% CI: 0.001-0.005, P<0.0001) and a mean direct effect of 0.023 (95% CI: 0.005-0.042, P=0.0060). the proportion of total effects mediated by the UHR was 12.28% (95% CI: 4.24%-43.13%, P=0.0020). These findings suggest a modest but significant role for UHR in the pathway linking sedentary time to hypertension.

## Discussion

This study utilized NHANES 2009-2018 data and found a significant positive correlation between sedentary time and hypertension, with this association partially mediated by the uric acid to high-density lipoprotein ratio (UHR), with a mediation proportion of 12.3%. Even after adjusting for confounding factors such as gender, age, race, and BMI, this relationship remained significant. The innovation of this study lies in the first identification of UHR as a mediator in the relationship between sedentary time and hypertension, providing a new perspective for exploring the potential mechanisms by which sedentary behavior leads to hypertension and suggesting that targeting UHR may be a potential strategy to reduce the risk of sedentary-related hypertension.

The umbrella review systematically assessed the effects of sedentary behavior on blood pressure and cardiovascular disease (CVD) incidence and mortality among adults aged 18 and older, including specific subgroups such as South Asians, stroke patients, and individuals with type 2 diabetes. The review analyzed 40 systematic reviews and meta-analyses, encompassing 393 individual studies with over 10 million participants. Previous studies, like those by Rezende et al., have highlighted the significant health impacts of sedentary behavior, noting associations with increased risks for mortality and cardiovascular events due to prolonged inactivity[12].

Specifically, the findings indicated a significant association between increased sedentary behavior and higher CVD incidence and mortality; decreased sedentary time was linked to lower CVD risk in 17 of 23 studies, supporting evidence presented by Dey et al. that suggests elevated sedentary behavior within South Asian populations could increase their CVD mortality risk[13] .However, the relationship between sedentary behavior and blood pressure was less consistent. Only 7 of 20 studies found that reduced sedentary time significantly lowered blood pressure, with interventional studies often demonstrating no noteworthy effects [13]. In contrast, Kanaya et al. noted that ethnic variances, such as those found in South Asians, contribute to different cardiovascular risk profiles, including hypertension, thereby complicating the direct effects of sedentary time on blood pressure across diverse populations[14].

Cross-sectional studies more frequently indicated negative impacts of sedentary behavior on blood pressure, which is consistent with findings from Kanaya et al., who observed that South Asians face a higher prevalence of diabetes, further exacerbating cardiovascular risks[14].Moreover, studies examining hypertension incidence predominantly indicated a detrimental effect of sedentary behavior, with an increased prevalence documented among South Asians[15]. This robust methodology underscored the substantial influence that sedentary lifestyles exert on cardiovascular health.[16].These findings underscore the complex interplay of sedentary time with cardiovascular diseases, though the relationship with hypertension remains ambiguous and appears significantly influenced by racial and ethnic factors. The evolving understanding of these relationships calls for further exploration, particularly in diverse demographic cohorts, to unravel the underlying mechanisms that contribute to cardiovascular risk associated with sedentary behavior.Our study is the first to discover the potential association between sedentary time and hypertension in the Nhanes population, especially among Mexican Americans. At the same time, it was found that UHR is involved in and regulates the association between sedentary time and the pathogenesis of hypertension.

The uric acid to high-density lipoprotein cholesterol ratio (UHR) is emerging as a valuable biomarker for cardiovascular and metabolic risk assessment [17,18]. Routine UHR monitoring, combined with hypertension risk factor management, enables early detection of metabolic dysregulation and hypertension. Personalized strategies, incorporating lifestyle, family history, and other risk factors, may enhance hypertension prevention [19]. Clinical guidelines should prioritize UHR measurement, timely intervention for abnormalities, and correction of uric acid and lipid imbalances. For elevated UHR, dietary adjustments (e.g., reducing high-purine foods, increasing antioxidant-rich produce) and moderate aerobic exercise are recommended to improve metabolic health and blood pressure control. Regular follow-up can further mitigate hypertension incidence in high-risk populations [20].

This study aims to utilize a large sample of NHANES 2017–2020 data to investigate the association between sedentary time and hypertension, while also examining the potential mediating role of the uric acid to high-density lipoprotein cholesterol ratio (UHR). The specific mechanisms by which sedentary behavior influences the pathogenesis and progression of hypertension through UHR remain not fully delineated. Current evidence suggests that visceral fat accumulation, insulin resistance, and disrupted uric acid metabolism may be critical factors in assessing the susceptibility of sedentary individuals to hypertension[21,22].Increased sedentary behavior is posited to slow metabolic activity, which may enhance the production of uric acid and simultaneously lower HDL levels, thereby establishing a state of elevated UHR.[23]. This high UHR state is thought to promote oxidative stress and inflammatory responses, which impair vascular endothelial function—a key event in the development of hypertension[24]. Moreover, elevated uric acid levels may activate the renin–angiotensin system, leading to vasoconstriction and sodium retention, while reduced HDL levels can diminish anti-inflammatory and antioxidative defenses; both effects contribute to endothelial dysfunction and elevated blood pressure[25-27].Furthermore, an increased UHR might interfere with insulin signaling pathways, thereby exacerbating insulin resistance and indirectly disrupting blood pressure regulation.[28] In individuals exhibiting high UHR, endothelial cell impairment is more pronounced, which compromises vasodilatory capacity and contributes to hypertension[29].

The results of this study provide substantial evidence for the correlation between sedentary time and hypertension, and the ratio of uric acid to high-density lipoprotein (UHR) plays a mediating role in this relationship. Since UHR can be obtained through routine blood tests, it is feasible to evaluate UHR in clinical practice. These results suggest that healthcare providers should not ignore the role of UHR when focusing on the impact of sedentary behavior on hypertension.

Regulating UHR levels may be an effective strategy to reduce the risk of hypertension related to sedentary behavior.

## Study strengths and limitations

This research has several significant advantages. Firstly, this is the first time that the NHANES large sample data system has been used to explore the relationship between sedentary duration and hypertension, and it has revealed part of the mediating role of uric acid to high-density lipoprotein ratio (UHR), providing a new perspective on the potential mechanism of sedentary behavior and hypertension. Secondly, the study adopted a multivariate adjustment model to ensure the robustness of the results, and verified the stability of the association through sensitivity analysis, enhancing the reliability of the conclusion. Furthermore, this study emphasizes the significance of UHR as a metabolic marker in sedentary hypertension, providing a potential target for clinical intervention.However, this study also has certain limitations. Due to the cross-sectional nature of the NHANES data, the causal relationship between sedentary time and hypertension cannot be determined. Prospective cohort studies are needed for further verification in the future. Secondly, the assessment of sedentary time relies on self-reported questionnaires, which may involve recall bias or measurement errors. Furthermore, although the study adjusted for confounding factors such as gender, age, race and BMI, there may still be unmeasured variables (such as diet and genetic factors) influencing the results. Finally, the biological mechanism of UHR still requires more experimental studies for in-depth exploration.

## Conclusion

In conclusion, this large-scale cross-sectional study indicates that prolonged sitting time is significantly associated with elevated UHR, and hypertension plays an important mediating role in the pathogenesis of both. This finding suggests that while public health intervention and clinical management reduce sedentary behavior, they should closely monitor the regulation of UHR levels. The synergistic intervention of the two may be an effective strategy to reduce the risk of hypertension.

## Data Availability

Data is provided within the manuscript or supplementary information files. Publicly available data were analyzed in this study, which can be found here: https://www.cdc.gov/nchs/nhanes/index.htm.

## Declarations

Ethics approval and consent to participate: The NHANES study protocol was approved by the National Center for Health Statistics (NCHS) Research Ethics Review Board. All participants provided written informed consent. The analysis of de-identified public data was exempt from additional ethical approval.

## Consent for publication

Not applicable. Clinical trial registration: Not applicable.

## Authors’ contributions

GL,Li, C,Wang: conceptualization. GL,Li: methodology and data curation. J,Tao: software.

GL,Li, C,Wang: validation.GL,Li, J,Fu: writing – original draft preparation.

GL,Li, J,Tao,Wang: writing – review and editing.GL,Li : visualization. All authors have read and agreed to the published version of the manuscript.

